# Asthma diagnosis in school-age children: a survey among Swiss physicians

**DOI:** 10.64898/2026.07.13.26357724

**Authors:** Beatriz Guerra Buezo, Mari Sasaki, Lorenz Manuel Leuenberger, Sarah Glick, Erol A Gaillard, Alexander Moeller, Nicolas Regamey, Oliver Sutter, Myrofora Goutaki, Claudia Elisabeth Kuehni

## Abstract

**Background:** Clinical guidelines recommend objective tests to diagnose asthma in school-age children, but their availability and use in routine practice remain uncertain. We evaluated asthma diagnostic practices in Switzerland, focusing on first-line tests (spirometry, bronchodilator reversibility testing, and fractional exhaled nitric oxide [FeNO]).

**Methods:** Cross-sectional, nationwide online survey of primary care paediatricians (PCPs) and respiratory specialists. We assessed access to and use of diagnostic tests, focusing on first-line tests, and examined reasons for non-use, referral practices, and guideline consultation. We used multivariable logistic regression to identified factors associated with spirometry access among PCPs.

**Results:** Of 1,055 respondents, 625 diagnosed asthma in children, including 419 PCPs. Among PCPs, 50% (95% confidence interval [CI] 45–55) reported no access to spirometry and 95% (95% CI 92–97) no access to FeNO, whereas all paediatric respiratory specialists and almost all adult respiratory specialists had access to both tests. Barriers to first-line testing among PCPs included economic constraints and difficulties interpreting test results. Spirometry access was lower in French– and Italian-speaking regions than in German-speaking regions (adjusted odds ratio [aOR] 0.09, 95% CI 0.05–0.15), but higher among PCPs working full-time (aOR 2.04, 95% CI 1.11–3.82) and those using Swiss asthma guidelines (aOR 1.71, 95% CI 1.01– 2.92). PCPs without spirometry access more frequently referred children to specialists for diagnostic confirmation (89% versus 80%; p=0.019).

**Conclusion:** Many PCPs in Switzerland lack access to guideline-recommended tests. Improving access, reimbursement, and training in test interpretation may help reduce the gap between guidelines and clinical practice.

## INTRODUCTION

Diagnosing asthma in children is challenging as the clinical presentation is heterogeneous, symptoms are unspecific and vary over time, and there is no single universally accepted diagnostic test [1–4]. Misdiagnosis leads to inappropriate treatment, uncontrolled symptoms, and delayed recognition of alternative conditions [5–7]. Therefore, recent guidelines emphasise the importance of confirming a clinical suspicion of asthma with objective testing. The European Respiratory Society (ERS) guideline for the diagnosis of asthma in school-age children [4] propose spirometry with bronchodilator reversibility testing (BDR) and fractional exhaled nitric oxide (FeNO) as first-line tests in the diagnostic work-up of suspected asthma. The Swiss recommendations are based on the ERS guidelines, but place more emphasis on user-friendliness [8].

In Switzerland, different physicians diagnose asthma in children: primary care paediatricians (PCPs), pulmonologists, allergologists and general practitioners. As all children with asthma are initially seen in primary care, PCPs play a central role in the diagnostic process [9–11]. In primary care settings, differences in training and access to objective tests may result in variation in diagnostic practices. Previous studies suggest asthma is commonly misdiagnosed in school-age children, particularly when diagnosis relies on history and symptoms without objective testing [12, 13]. For example, a Dutch primary care study from 2016, including 652 children aged 6–18 years, found that only 16% had asthma confirmed by spirometry, while more than half of those previously labelled with asthma were likely overdiagnosed [14]. Similarly, a Canadian community-based study reported that only 18% of children with physician-diagnosed asthma had undergone pulmonary function testing, and up to 45% were misdiagnosed [6]. Representative studies conducted after the publication of the ERS guideline are lacking.

In 2024, three years after publication of the ERS guideline [4] and a few months after the simplified Swiss guidelines [8], we conducted a nationwide survey of physicians involved in diagnosing asthma in school-age children at primary, secondary and tertiary care levels in Switzerland, with a particular focus on PCPs. We describe the access to asthma diagnostic tests in clinical practice and barriers to using first-line tests. We also describe physician characteristics associated with access to spirometry, a key first-line diagnostic test, and examine how access to diagnostic tests influences referrals to specialists.

## METHODS

### Questionnaire design

We developed the initial questionnaire in English based on expert consensus, piloted it among PCPs and respiratory specialists, and revised it. The final questionnaire was translated by native speakers into German, French, and Italian, the main official languages in Switzerland and piloted again. It consisted of 50 core questions with conditional branching, divided into five sections **(Table S1)**: (1) information on the demographics and clinical practice of physicians, (2) access and use of asthma diagnostic tests, (3) follow-up and referral practices, (4) use of a medication trial for asthma diagnosis, and (5) guideline consultation. Conditional branching allowed to tailor questions according to specialty and access to tests. Only physicians seeing school-age children (5-16 years) with suspected asthma completed all sections.

### Study design and data collection

From May to August 2024, we conducted an anonymous, cross-sectional online survey of physicians involved in diagnosing asthma in school children in Switzerland. We contacted participants through four medical societies: the Swiss Paediatric Society, the Swiss Society of Paediatric Pulmonology, the Swiss Society of Pulmonology, and the Swiss Society for Allergology and Immunology. Questionnaire distribution differed between societies. Members of the Swiss Paediatric Society and the Swiss Society of Paediatric Pulmonology received personalised email invitations followed by three personalised reminders at intervals of three weeks. The Swiss Society of Pulmonology and the Swiss Society for Allergology and Immunology both distributed the survey link to their full membership via email, with one general reminder after one month.

For the analysis, we classified respondents into five groups according to their specialty and workplace: 1) primary care paediatricians (PCPs), defined as general paediatricians working in a non-hospital outpatient practice; 2) hospital paediatricians, defined as general paediatricians working in a hospital without subspecialty certification; 3) paediatric asthma specialists, defined as paediatric pulmonologists and paediatric allergologists); 4) adult asthma specialists, defined as adult pulmonologists and adult allergologists; and 5) others, defined as paediatricians with a subspecialty other than pulmonology or allergology. Results for this group are not reported, as these physicians might see a child with asthma on the ward, but are not involved in the diagnostic process.

We used Research Electronic Data Capture (REDCap) to collect and manage data from the survey [15]. Under Swiss legislation, we were not required to obtain ethics committee approval or informed consent because the study was addressed to physicians and did not involve health-related personal data.

### Statistical analysis

We analysed data from all physicians who reported evaluating school-age children with suspected asthma and who completed the full survey. We excluded other paediatric specialists because they were not involved in the diagnosis process. We summarised participant characteristics and access and use of diagnostic tests using counts and percentages with 95% confidence intervals (CI) for categorical variables. For questions allowing multiple responses, we calculated proportions separately for each response option. We examined factors associated with access to spirometry in primary care using univariable and multivariable logistic regression restricted to PCPs. The dependent variable was self-reported access to spirometry (access versus no access). Independent variables included respondent gender, age group, language region, clinical workload (full-time versus part-time), number of children with asthma seen per month, number of new asthma diagnoses per month, and consultation of clinical guidelines. Multivariable models included all listed variables, and results are presented as adjusted odds ratios (aORs) with 95% CI. We conducted all analyses using R (version 4.4.1) [16]. We report this study in accordance with the Strengthening the Reporting of Observational Studies in Epidemiology (STROBE) guidelines for cross-sectional studies [15].

## RESULTS

The response rate varied substantially between medical societies **(Figure S1)**. It reached 42% in the Swiss Paediatric Society (885/2,130 members) and 81% (51/63) among paediatric pulmonologists. In contrast, only 14% (81/575) of adult pulmonologists and 6% (14/226) of allergologists answered. Twenty-two physicians were members of both the Swiss Paediatric Society and the Swiss Society of Paediatric Pulmonology, and we allocated them to the Swiss Society of Paediatric Pulmonology.

Respondents consisted of 419 PCPs, 109 hospital paediatricians, 56 paediatric asthma specialists and 41 adult asthma specialists **(Table 1).** Among participating PCPs, 63% reported German as their main working language. Respondents were predominantly female (75%). Most PCPs reported seeing 1-5 children with asthma per month. Regarding guideline use, 92% of PCPs reported consulting clinical guidelines for asthma diagnosis. Across groups, physicians most often used the Swiss 2024 asthma recommendations [8]. Paediatric asthma specialists reported the highest use of the ERS 2021 guideline [4], whereas adult asthma specialists more frequently consulted the Global Initiative for Asthma (GINA) 2023 guideline [17] **(Table 2).**

**Table 1.**
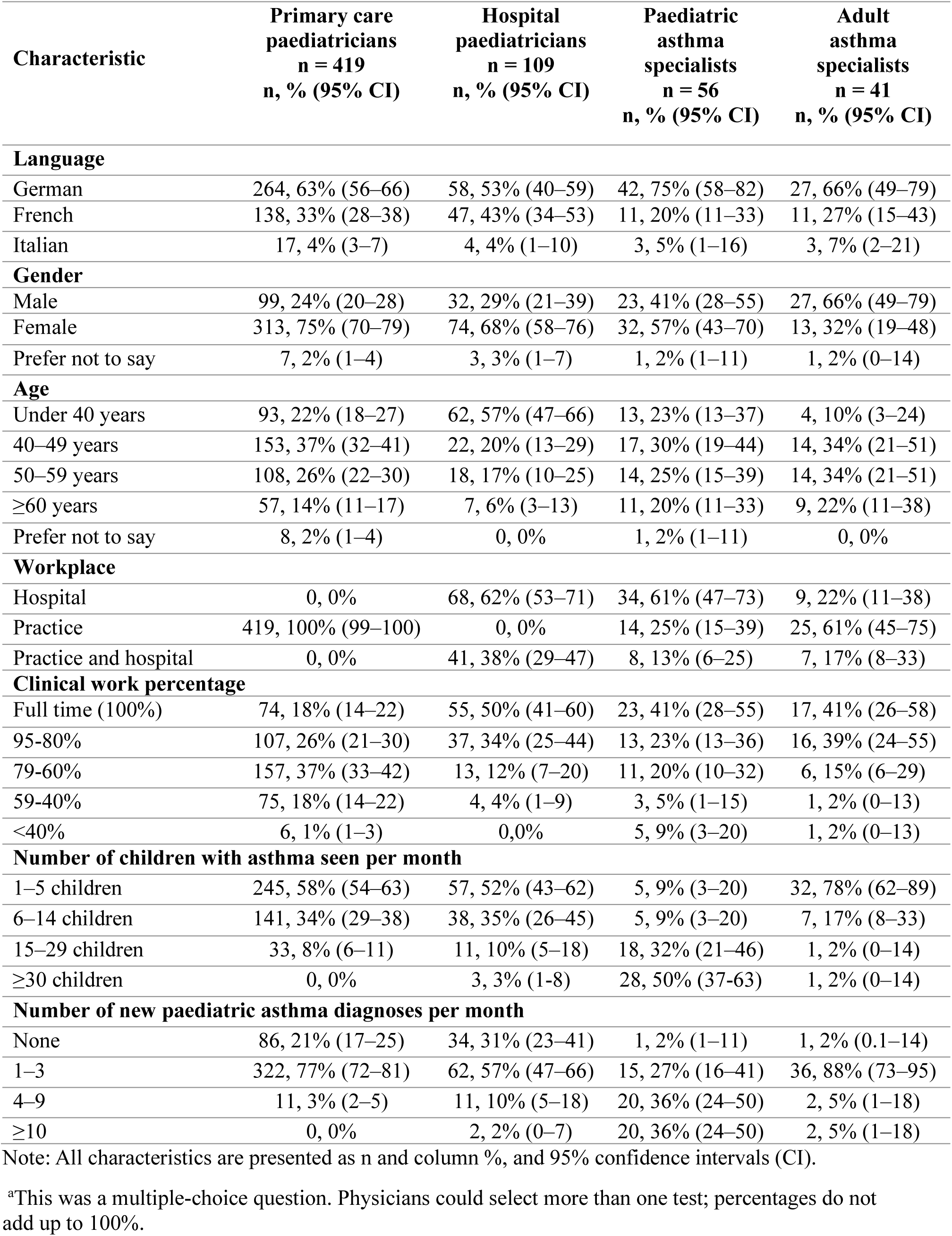
Demographic and clinical practice characteristics of the Swiss physicians participating in the survey, grouped by physician type: primary care paediatricians, hospital paediatricians, paediatric specialists and adult specialists (n=625).

| Characteristic | Primary care paediatricians<br>n = 419<br>n, % (95% CI) | Hospital paediatricians<br>n = 109<br>n, % (95% CI) | Paediatric asthma specialists<br>n = 56<br>n, % (95% CI) | Adult asthma specialists<br>n = 41<br>n, % (95% CI) |
| --- | --- | --- | --- | --- |
| **Language** |  |  |  |  |
| German | 264, 63% (56–66) | 58, 53% (40–59) | 42, 75% (58–82) | 27, 66% (49–79) |
| French | 138, 33% (28–38) | 47, 43% (34–53) | 11, 20% (11–33) | 11, 27% (15–43) |
| Italian | 17, 4% (3–7) | 4, 4% (1–10) | 3, 5% (1–16) | 3, 7% (2–21) |
| **Gender** |  |  |  |  |
| Male | 99, 24% (20–28) | 32, 29% (21–39) | 23, 41% (28–55) | 27, 66% (49–79) |
| Female | 313, 75% (70–79) | 74, 68% (58–76) | 32, 57% (43–70) | 13, 32% (19–48) |
| Prefer not to say | 7, 2% (1–4) | 3, 3% (1–7) | 1, 2% (1–11) | 1, 2% (0–14) |
| **Age** |  |  |  |  |
| Under 40 years | 93, 22% (18–27) | 62, 57% (47–66) | 13, 23% (13–37) | 4, 10% (3–24) |
| 40–49 years | 153, 37% (32–41) | 22, 20% (13–29) | 17, 30% (19–44) | 14, 34% (21–51) |
| 50–59 years | 108, 26% (22–30) | 18, 17% (10–25) | 14, 25% (15–39) | 14, 34% (21–51) |
| ≥60 years | 57, 14% (11–17) | 7, 6% (3–13) | 11, 20% (11–33) | 9, 22% (11–38) |
| Prefer not to say | 8, 2% (1–4) | 0, 0% | 1, 2% (1–11) | 0, 0% |
| **Workplace** |  |  |  |  |
| Hospital | 0, 0% | 68, 62% (53–71) | 34, 61% (47–73) | 9, 22% (11–38) |
| Practice | 419, 100% (99–100) | 0, 0% | 14, 25% (15–39) | 25, 61% (45–75) |
| Practice and hospital | 0, 0% | 41, 38% (29–47) | 8, 13% (6–25) | 7, 17% (8–33) |
| **Clinical work percentage** |  |  |  |  |
| Full time (100%) | 74, 18% (14–22) | 55, 50% (41–60) | 23, 41% (28–55) | 17, 41% (26–58) |
| 95–80% | 107, 26% (21–30) | 37, 34% (25–44) | 13, 23% (13–36) | 16, 39% (24–55) |
| 79–60% | 157, 37% (33–42) | 13, 12% (7–20) | 11, 20% (10–32) | 6, 15% (6–29) |
| 59–40% | 75, 18% (14–22) | 4, 4% (1–9) | 3, 5% (1–15) | 1, 2% (0–13) |
| <40% | 6, 1% (1–3) | 0, 0% | 5, 9% (3–20) | 1, 2% (0–13) |
| **Number of children with asthma seen per month** |  |  |  |  |
| 1–5 children | 245, 58% (54–63) | 57, 52% (43–62) | 5, 9% (3–20) | 32, 78% (62–89) |
| 6–14 children | 141, 34% (29–38) | 38, 35% (26–45) | 5, 9% (3–20) | 7, 17% (8–33) |
| 15–29 children | 33, 8% (6–11) | 11, 10% (5–18) | 18, 32% (21–46) | 1, 2% (0–14) |
| ≥30 children | 0, 0% | 3, 3% (1–8) | 28, 50% (37–63) | 1, 2% (0–14) |
| **Number of new paediatric asthma diagnoses per month** |  |  |  |  |
| None | 86, 21% (17–25) | 34, 31% (23–41) | 1, 2% (1–11) | 1, 2% (0.1–14) |
| 1–3 | 322, 77% (72–81) | 62, 57% (47–66) | 15, 27% (16–41) | 36, 88% (73–95) |
| 4–9 | 11, 3% (2–5) | 11, 10% (5–18) | 20, 36% (24–50) | 2, 5% (1–18) |
| ≥10 | 0, 0% | 2, 2% (0–7) | 20, 36% (24–50) | 2, 5% (1–18) |

Note: All characteristics are presented as n and column %, and 95% confidence intervals (CI).
<sup>a</sup>This was a multiple-choice question. Physicians could select more than one test; percentages do not add up to 100%.

**Table 2.** Guideline use of the Swiss physicians participating in the survey, grouped by physician type: primary care paediatricians, hospital paediatricians, paediatric specialists, and adult specialists.

| Guideline use | Primary care paediatricians<br>n = 419<br>n, % (95% CI) | Hospital paediatricians<br>n = 109<br>n, % (95% CI) | Paediatric asthma specialists<br>n = 56<br>n, % (95% CI) | Adult asthma specialists<br>n = 41<br>n, % (95% CI) |
| --- | --- | --- | --- | --- |
| **Do you consult any clinical guidelines?** |  |  |  |  |
| Yes | 384, 92 % (89-94) | 101, 93% (88-98) | 56, 100% | 38, 93% (85-98) |
| No | 35, 8% (6-11) | 8, 7% (2-12) | 0, 0% | 3, 7% (0-15) |
| **Which guideline do you consult?** (Multiple choice question)<sup>a</sup> |  |  |  |  |
| ERS guideline 2021<sup>b</sup> | 37, 10% (7-13) | 14, 14% (8-22) | 35, 62% (49-74) | 15, 39% (26-55) |
| Swiss guideline 2024<sup>c</sup> | 295, 77% (72-81) | 76, 75% (66-83) | 50, 89% (79-95) | 23, 61% (45-74) |
| GINA guideline 2023<sup>d</sup> | 193, 50% (45-55) | 58, 57% (48-67) | 33, 59% (46-71) | 28, 74% (58-85) |
| Swiss guideline 2009<sup>e</sup> | 43, 11% (8-15) | 10, 10% (5-17) | 2, 4% (1-12) | 6, 16%; (7-30) |
| German guidelines 2017<sup>f</sup> | 9, 2% (1-4) | 2, 2% (1-7) | 4, 7% (3-17) | 6, 16% (7-30) |
| French guidelines 2022<sup>g</sup> | 11, 3% (2-5) | 2, 2% (1-7) | 2, 4% (1-12) | 1, 3% (0-13) |
| Another guideline | 2, 1% (0-2) | 1, 1% (0-5) | 1, 2% (0-9) | 1, 3% (0-13) |

Note: Percentages and 95% confidence intervals (CI).
<sup>a</sup> Multiple choice question, respondents could select multiple reasons. Percentages are based on the number of physicians who reported using the clinical guidelines.
<sup>b</sup> European Respiratory Society clinical practice guideline for the diagnosis of asthma in children aged 5–16 years (Gaillard et al., 2021)
<sup>c</sup> Swiss recommendations for the diagnosis, treatment, and management of asthma in children aged ≥5 years (Moeller et al., 2024).
<sup>d</sup> Global Strategy for Asthma Management and Prevention, Global Initiative for Asthma (GINA) Report (Global Initiative for Asthma, 2023).
<sup>e</sup> Swiss national recommendations for the diagnosis and management of asthma in children (Roth et al., 2009).
<sup>f</sup> Guideline of the German Respiratory Society for the diagnosis and management of asthma (Buhl et al., 2017).
<sup>g</sup> Guidelines for management and follow-up of asthmatic patients of the French Society of Pneumology and the French Society of Pediatric Pneumology and Allergology. (Raheison-Semjen et al., 2022).

### Access and use of asthma diagnostic tests

Among all PCPs, 129 (31%) used spirometry in almost all children (> 90%) with suspected asthma and 47 (11%) used it in most (50-90%). Half of PCPs reported no access to spirometry in their practice. Among PCPs with access to spirometry, only 81 (19%) reported performing BDR in almost all children. Only 11 PCPs (3%) used FeNO in almost all children and 5 (1%) used it in most. FeNO was not available to 398 (95%) of PCPs. In contrast to PCPs, nearly all paediatric asthma specialists reported access to spirometry, BDR and FeNO, and used these tests routinely in the diagnostic assessment of children with suspected asthma. Adult asthma specialists also reported high access to first-line tests, although use of FeNO and spirometry was a bit lower. Nearly two thirds of adult and paediatric asthma specialists used also whole-body plethysmography in all or most children. Allergy testing was available in most practices, with only 55 (13%) PCPs reporting no access. Among all PCPs, 205 (49%) used allergy testing in almost all children and 80 (19%) used it in most. This proportion was higher among paediatric asthma specialists, 51 (91%) of whom reported using allergy testing in almost all children. PCPs rarely or never used peak expiratory flow measurements or chest X-rays to diagnose asthma **(Table 3).**

**Table 3.** Access and use of diagnostic tests for asthma diagnosis of school-age children.

|  | **Primary care**<br>**paediatricians**<br>**n = 419**<br>**n, % (95% CI)** | **Hospital**<br>**paediatricians**<br>**n = 109**<br>**n, % (95% CI)** | **Paediatric asthma**<br>**specialists**<br>**n = 56**<br>**n, % (95% CI)** | **Adult asthma**<br>**specialists**<br>**n = 41**<br>**n, % (95% CI)** |
| --- | --- | --- | --- | --- |
| **Spirometry** |  |  |  |  |
| Used in almost all (>90%) | 129, 31% (26-35) | 28, 26% (18-35) | 55, 98% (90-100) | 37, 90% (77-97) |
| Used in most (50-90%) | 47, 11% (8-15) | 7, 6% (3-13) | 1, 2% (0-10) | 2, 5% (1-17) |
| Infrequently used (<50%) | 34, 8% (6-11) | 10, 9% (4-16) | 0, 0% | 0, 0% |
| Available but never used | 8, 2% (1-4) | 10, 9% (4-16) | 0, 0% | 0, 0% |
| Not available | 201, 48% (43-53) | 54, 49% (40-59) | 0, 0% | 2, 5% (1-17) |
| **Bronchodilator response (BDR)<sup>a</sup>** |  |  |  |  |
| Used in almost all (>90%) | 81, 19% (16-23) | 19, 17% (11-26) | 48, 86% (74-93) | 29, 71% (55-84) |
| Used in most (50-90%) | 43, 10% (7-14) | 8, 7% (3-14) | 7, 12% (5-24) | 8, 19% (9-35) |
| Infrequently used (<50%) | 46, 11% (8-14) | 4, 4% (1-9) | 1, 2% (0-10) | 2, 5% (1-17) |
| Not available | 249, 59% (55-64) | 78, 72% (62-80) | 0, 0% | 2, 5% (1-17) |
| **FeNO** |  |  |  |  |
| Used in almost all (>90%) | 11, 3% (1-5) | 19, 17% (11-26) | 55, 98% (90-100) | 35, 85% (71-94) |
| Used in most (50-90%) | 5, 1% (0-3) | 1, 1% (0-5) | 1, 2% (0-10) | 2, 5% (1-17) |
| Infrequently used (<50%) | 5, 1% (0-2) | 10, 9% (4-16) | 0, 0% | 1, 2% (0-13) |
| Not available | 398, 95% (92-97) | 79, 72% (63-81) | 0, 0% | 3, 7% (2-20) |
| **Peak expiratory flow (PEF)** |  |  |  |  |
| Used in almost all (>90%) | 16, 4% (2-6) | 3, 3% (1-8) | 0, 0% | 5, 12% (4-26) |
| Used in most (50-90%) | 9, 2% (1-4) | 0, 0% | 0, 0% | 2, 5% (1-17) |
| Infrequently used (<50%) | 35, 8% (6-11) | 8, 7% (3-14) | 4, 7% (2-17) | 21, 51% (35-67) |
| Not available | 359, 86% (82-89) | 98, 90% (83-95) | 52, 93% (83-98) | 13, 32% (18-48) |
| **Allergy testing** |  |  |  |  |
| Used in almost all (>90%) | 205, 49% (44-54) | 37, 34% (25-44) | 51, 91% (80-97) | 21, 51% (35-67) |
| Used in most (50-90%) | 80, 19% (15-23) | 14, 13% (7-21) | 5, 9% (3-20) | 8, 19% (9-35) |
| Infrequently used (<50%) | 79, 19% (15-23) | 13, 12% (7-19) | 0, 0% | 8, 19% (9-35) |
| Not available | 55, 13% (10-17) | 45, 41% (32-51) | 0, 0% | 4, 10% (3-23) |
| **Whole-body plethysmography** |  |  |  |  |
| Used in almost all (>90%) | 2, 0% (0-2) | 8, 7% (3-14) | 25, 45% (31-59) | 26, 63% (47-78) |
| Used in most (50-90%) | 0, 0% | 9, 8% (4-15) | 9, 16% (8-28) | - |
| Infrequently used (<50%) | 3, 1% (0-2) | 3, 3% (1-8) | 8, 14% (6-26) | 3, 7% (2-20) |
| Not available | 414, 99% (97-99) | 89, 81% (73-89) | 14, 25% (10-50) | 12, 29% (13-53) |
| **Chest X-ray** |  |  |  |  |
| Used in almost all (>90%) | 5, 1% (0-3) | 4, 4% (1-9) | 1, 2% (0-10) | 0, 0% |
| Used in most (50-90%) | 8, 2% (1-4) | 4, 4% (1-9) | 3, 5% (1-15) | 0, 0% |
| Infrequently used (<50%) | 64, 15% (12-19) | 29, 27% (19-36) | 8, 14% (6-26) | 7, 17% (7-32) |
| Not used for diagnosis | 342, 82% (78-85) | 72, 66% (56-75) | 44, 79% (66-88) | 34, 83% (68-93) |

Abbreviations: FeNO, fractional exhaled nitric oxide. Note: All characteristics are presented as n and column %, and 95% confidence intervals (CI). <sup>a</sup>The use of BDR was only assessed physicians who reported access to spirometry. Physicians without a spirometer were categorized as “not available” for the use of BDR.

### Barriers for access to first line tests among primary care paediatricians

PCPs without access to spirometry (n = 208) or FeNO (n = 398) reported multiple reasons **(Table 4).** For spirometry, commonly reported organisational barriers included lack of staff (38%) and limited consultation time (36%), while economic barriers included device costs (28%) and inadequate reimbursement (14%). In addition, 41% of PCPs reported difficulties interpreting spirometry results in children. For FeNO, economic factors were the main barrier, including device costs (27%) and reimbursement (21%). Organisational barriers, such as lack of time or staff, were reported by 19% of PCPs, while 26% indicated difficulty interpreting FeNO results. One in five paediatricians indicated that they did not consider spirometry or FeNO necessary for asthma diagnosis. Free-text explanations given by PCPs further explain stated barriers **(Table S2).**

**Table 4.** Barriers to access to first-line tests for asthma diagnosis in school-age children among PCPs.

| **Reasons for not having these tests** | **Spirometry**<br>**n = 208**<br>**n, (%)** | **FeNO**<br>**n = 398**<br>**n, (%)** |
| --- | --- | --- |
| **Clinical reasons** |  |  |
| I don't need this test for asthma diagnosis | 56 (20) | 81 (20) |
| I refer children to specialists | 11 (5) | 29 (28) |
| **Economic reasons** |  |  |
| I find the device too expensive | 56 (28) | 118 (27) |
| I am reimbursed poorly for the test | 29 (14) | 83 (21) |
| **Organisational barriers** |  |  |
| I don't have enough staff | 77 (38) | 79 (20) |
| I don't have enough time | 99 (36) | 97 (19) |
| **Test-related challenges** |  |  |
| I find the test difficult to perform with children | 18 (9) | 6 (2) |
| I find the results difficult to interpret in children | 84 (41) | 104 (26) |
| **Other reasons** |  |  |
| Other<sup>a</sup> (Free text responses; for examples, see table S3) | 22 (11) | 57 (32) |

*Abbreviations: FeNO, fractional exhaled nitric oxide*
Note: Multiple choice question, respondents could select multiple reasons. Percentages are based on the number of primary care paediatricians without access to each test. <sup>a</sup>Unique free-text responses that did not fit into other categories.

### Factors associated with spirometry access

We investigated factors associated with access to spirometry among PCPs **(Figure 1 and Table S3).** In multivariable logistic regression models, PCPs working in French– or Italian– speaking regions had much lower odds of having spirometry than those working in the German– speaking region (aOR 0.09, 95% CI 0.05–0.15). PCPs working full-time had higher odds of spirometry access than those working part-time (aOR 2.04, 95% CI 1.11–3.82). Use of the Swiss 2024 asthma recommendations was associated with higher spirometry access (aOR 1.71, 95% CI 1.01–2.92). Gender, age group, number of children with asthma seen per month, number of new asthma diagnoses made per month, and consultation of the ERS or GINA asthma diagnosis guidelines were not significantly associated with access to spirometry in the adjusted models. We could not investigate factors associated with access to FeNO, used by only 22 PCPs (5%).

**Figure 1.**
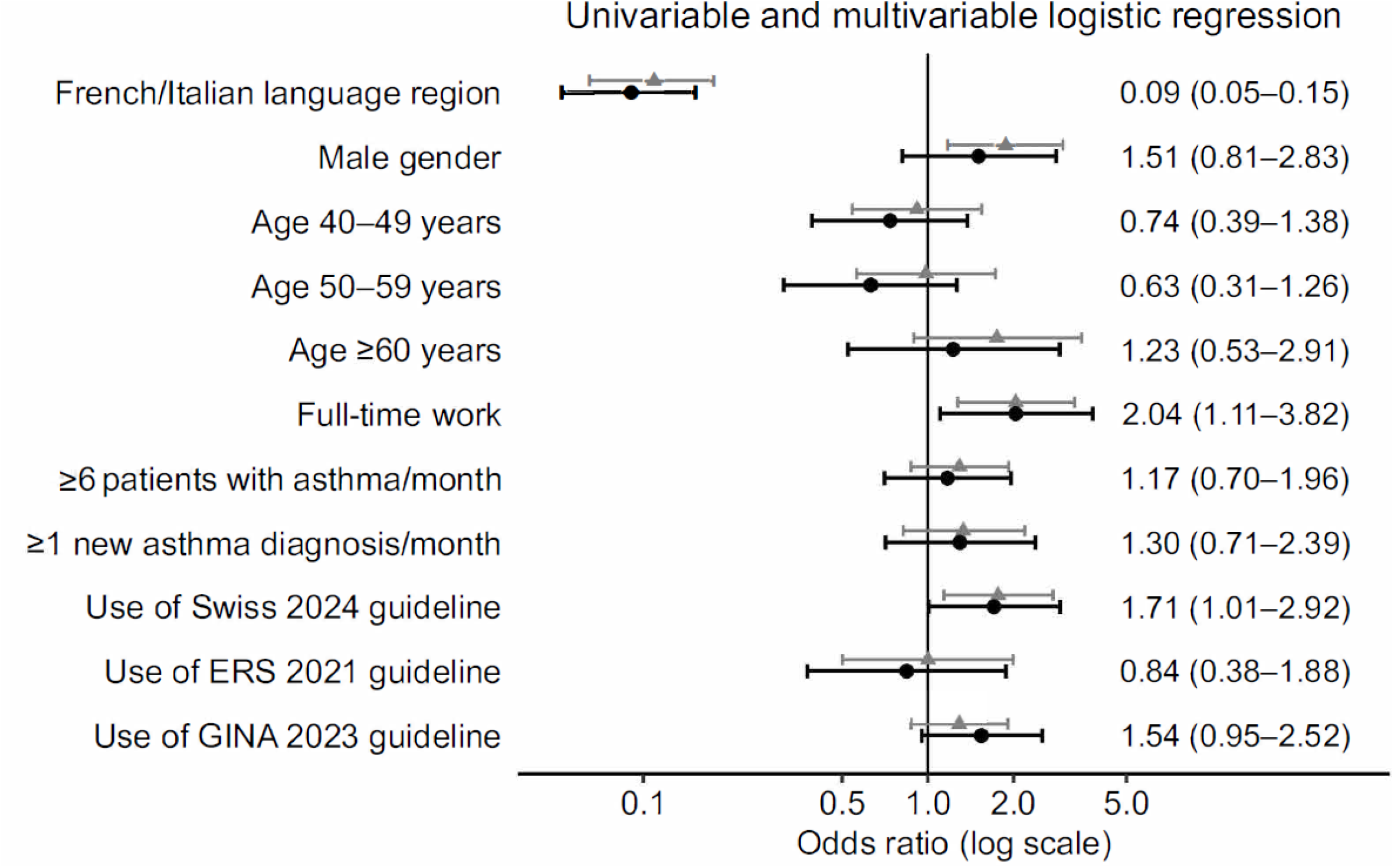
Factors associated with access to spirometry among primary care paediatricians when evaluating school-age children for asthma (n=419). Univariable and multivariable logistic regression. Note: Grey lines with triangles indicate unadjusted odds ratios (OR) and black lines with circles adjusted odds ratios (aOR). Horizontal lines represent 95% confidence intervals. Multivariable models were adjusted for gender, age group, language region, clinical workload, asthma caseload, number of new diagnoses, and guideline consultation. *Reference categories: German region, Female, <40 years, Part-time work percentage (<80%), 1–5 children with asthma seen per month, No new asthma diagnoses per month, No use of Swiss 2024, ERS 2021, or GINA 2023 guidelines. Odds ratios >1 indicate higher odds of using spirometry compared with the reference category*.

### Referrals to specialists among primary care paediatricians

Referral practices varied among PCPs: 114 (27%) reported referring more than 90% of children with suspected asthma to a specialist, 137 (33%) referred 50–90%, 112 (27%) referred 10–49%, and 56 (13%) referred fewer than 10% **(Table 5, Figure S2).** Among PCPs without access to spirometry, 179 (89%) reported referring children because diagnostic tests were unavailable in their practice, compared with 175 (80%) of those with access (p = 0.019). By contrast, PCPs with access to spirometry more often referred children for severe symptoms, long-term management, improving treatment adherence, managing comorbidities, or complex social or family situations.

**Table 5.** Referral reasons of children with suspected asthma to a specialist reported by primary care paediatricians (n = 419).

| Referral among primary care paediatricians | All PCP<br>n, (%)<br>419 (100) | PCP with<br>spirometry<br>n, (%)<br>218 (52) | PCP without<br>spirometry<br>n, (%)<br>201 (48) | p-value |
| --- | --- | --- | --- | --- |
| **What percentage of children do you refer to a specialist?** |  |  |  | <0.0001 |
| > 90 % | 114 (27) | 38 (17) | 76 (38) |  |
| 50-90 % | 137 (33) | 60 (28) | 77 (38) |  |
| 10-49 % | 112 (27) | 79 (36) | 33 (16) |  |
| < 10 % | 56 (13) | 41 (19) | 15 (8) |  |
| **Where do you usually refer your patients?** |  |  |  | <0.0001 |
| Specialist practice | 97 (23) | 38 (18) | 59 (30) |  |
| Specialist consultation at the hospital | 165 (40) | 105 (48) | 60 (30) |  |
| Depend on the case | 152 (36) | 74 (34) | 78 (39) |  |
| Not answered | 5 (1) | 1 (0) | 4 (1) |  |
| **I refer the children to a specialist...** |  |  |  |  |
| To perform tests that are not available to me | 354 (84) | 175 (80) | 179 (89) | 0.019 |
| To manage severe symptoms | 336 (80) | 188 (86) | 148 (74) | 0.002 |
| To confirm the diagnosis of asthma | 300 (71) | 147 (67) | 153 (76) | 0.063 |
| To comply with the family's request | 295 (70) | 170 (78) | 75 (62) | <0.0001 |
| To improve adherence to therapy | 190 (45) | 120 (55) | 70 (35) | <0.0001 |
| To manage cases with comorbidities | 187 (45) | 112 (51) | 75 (37) | 0.005 |
| To intensify the asthma therapy | 99 (23) | 39 (18) | 60 (30) | 0.006 |
| To deal with a complex or social family situation | 93 (22) | 62 (28) | 31 (15) | 0.002 |
| To reduce or discontinue the asthma therapy | 17 (4) | 6 (3) | 11 (6) | 0.245 |

<sup>a</sup>This was a multiple-choice question. Physicians could select more than one option; percentages do not add up to 100%.
Note: Data are presented as n (column %), unless otherwise stated. Percentages were calculated within each group of primary care paediatricians. P-values were calculated using Pearson's $\chi^2$ test, except for the variable *Percentage of children referred*, for which the Cochran–Armitage trend test was used.

## DISCUSSION

This nationwide survey, conducted three years after the publication of the ERS guideline for asthma diagnosis in school-age children [4] and a few months after publication of the closely related Swiss guidelines [8] provides updated evidence that access to and use of first-line diagnostic tests are excellent among specialists in Switzerland, but remain limited in primary care, affecting the implementation of asthma guidelines. Half of PCPs reported no access to spirometry, and almost all reported no access to FeNO. Barriers to accessing these tests were educational, organisational, and economic. Spirometry access varied strongly by region and was much lower in French– and Italian-speaking regions. Physicians working full– time and those consulting the Swiss national asthma recommendations had higher spirometry access.

### Interpretation of findings

A striking result is that FeNO was hardly available in primary care. In Switzerland, FeNO testing is currently not reimbursed by health insurance companies for general practitioners or general paediatricians without subspecialty certification in pulmonology or allergology, whereas spirometry is reimbursed without restriction. This difference might largely explain the marked disparity in availability between the two tests.

Regional differences in the use of spirometry may reflect variations in healthcare organisation between cantons in Switzerland, medical training traditions, proximity to specialised paediatric respiratory centres, and paediatrician density, which is higher in French– and Italian speaking regions, probably representing better access to specialist care [18]. We cannot further examine reasons for the discrepancy from our survey data. Previous surveys among physicians in Switzerland, on the management of acute bronchiolitis and on the use of complementary medicine among paediatricians, also described profound practice differences between German-speaking Switzerland and the French– and Italian-speaking regions [19, 20]. Better access to asthma diagnostic tests in primary care could reduce the need for specialist referrals and ensure these services are used more efficiently.

Our findings are consistent with previous international surveys reporting limited access of spirometry in paediatric primary care [21, 22]. A 2007 survey of 320 paediatricians and family physicians in the US found that only 21% of PCPs used spirometry for asthma diagnosis [23], while a 2009 European survey of 436 ambulatory paediatricians across six European countries (Belgium, France, Germany, Italy, Luxembourg and Slovenia) revealed that spirometry was available in fewer than 40% of paediatric primary care settings in most countries [24]. More recent data show minimal progress: a 2023 US survey found that spirometry remained underused, with only 16 of 37 paediatricians reporting access [25].

Despite increased awareness and clearer guideline recommendations, our findings show that spirometry remains under-utilized in paediatric primary care even in a high-income country like Switzerland. FeNO is recommended as a first-line diagnostic test in recent clinical guidelines [4, 8, 25], but there is little data on its availability and use in paediatric primary care. To our knowledge, our study is the first to systematically evaluate access to and use of FeNO in routine paediatric practice.

We identified several barriers in our study also reported elsewhere. A 2023 systematic review of Australian general practices identified insufficient training in spirometry interpretation, time constraints, and cost as the main barriers [26]. Similar challenges have been reported in other countries, suggesting they are not unique to Switzerland [23, 27, 28]. Awareness of clinical guidelines does not necessarily ensure their implementation. A 2018 study including paediatricians from Sweeden, Netherlands, Poland, Italy, Germany and Cyprus found that, although asthma guidelines were available in all countries, their implementation remained limited [29]. Similarly, a qualitative study of 30 Dutch general practitioners in 2009 identified organisational constraints, insufficient knowledge, and unclear recommendations as barriers to implementation [30].

The marked contrast between PCPs and asthma specialists suggests that implementation barriers are concentrated in primary care rather than reflecting a lack of availability of these tests within the Swiss healthcare system. However, barriers alone may not fully explain why first-line tests are not widely used. A key challenge of spirometry and FeNO is that they have a relatively low diagnostic yield in children, particularly in primary care settings where results are often normal or inconclusive [31]. Together, this may explain why adherence to asthma recommendations is often limited in routine clinical practice [29, 30, 32–35].

### Strengths and limitations

This is the first nationwide survey in Switzerland that examined access to and use of objective diagnostic tests for childhood asthma in paediatric primary care and among respiratory specialists. It was conducted after the publication of national and international asthma guidelines that provided clear recommendations on the use of Spirometry and FeNO as first-line tests for asthma diagnosis. Strengths of the study include its nationwide distribution, inclusion of physicians from different specialties and care settings, and the detailed assessment of diagnostic practices and perceived barriers.

We should consider several limitations. Firstly, while the response rate among paediatricians was good (42%), it does not exclude selection bias. Physicians with a particular interest in asthma diagnosis may have been more likely to participate, this may have led potentially to an overestimation of access to tests. Our findings mainly reflect paediatric practice and are not generalisable to all primary care settings, but over 80% of children under 18 in Switzerland have a paediatrician as their primary care provider. Also, response rates among adult specialists were low, but this may also be because few of them see children. Secondly, the study relied on self-reported data, which may be subject to recall bias or social desirability bias. Finally, we could not directly investigate factors such as training background and educational opportunities that may influence access to and use of diagnostic tests.

### Implications and recommendations

Our findings highlight a clear gap between guideline recommendations and real-world clinical practice in Switzerland. Improving access to first-line diagnostic tests in primary care is essential to support guideline-based asthma diagnosis, reduce reliance on clinical judgement alone, and minimise misdiagnosis and inappropriate management [4, 5, 8, 14, 17, 36]. However, improving access alone is insufficient. In our survey, 41% of PCPs reported difficulties interpreting results, indicating a need for further training. Educational strategies should focus on practical skills, including performing and interpreting lung function tests, rather than just disseminating guideline recommendations, as education alone rarely leads to lasting changes [37, 38]. In line with this, a 2016 UK study of paediatric primary care (10 general practices, 612 children) showed that, with practical training, staff were able to perform and interpret spirometry and FeNO in children [39].

In addition to training needs, our results point to important structural barriers. Financial constraints, including insufficient or – for FeNO – lack of reimbursement and lack of staff or time were frequently reported. Similar challenges have been described in the UK, where a 2025 report evaluating the BTS/NICE/SIGN asthma guideline identified insufficient resources and funding as key barriers to implementation [36, 40]. Closing the gap between guideline recommendations and clinical practice requires a combined approach integrating education, organisational support, and policy change. Finally, future guidelines may need to address how recommendations can be implemented in primary care and other settings, such as low– and middle-income countries or regions where patients have to travel long distances to see specialists, resulting in limited access to the recommended diagnostic tests (**Table 6**).

**Table 6.**
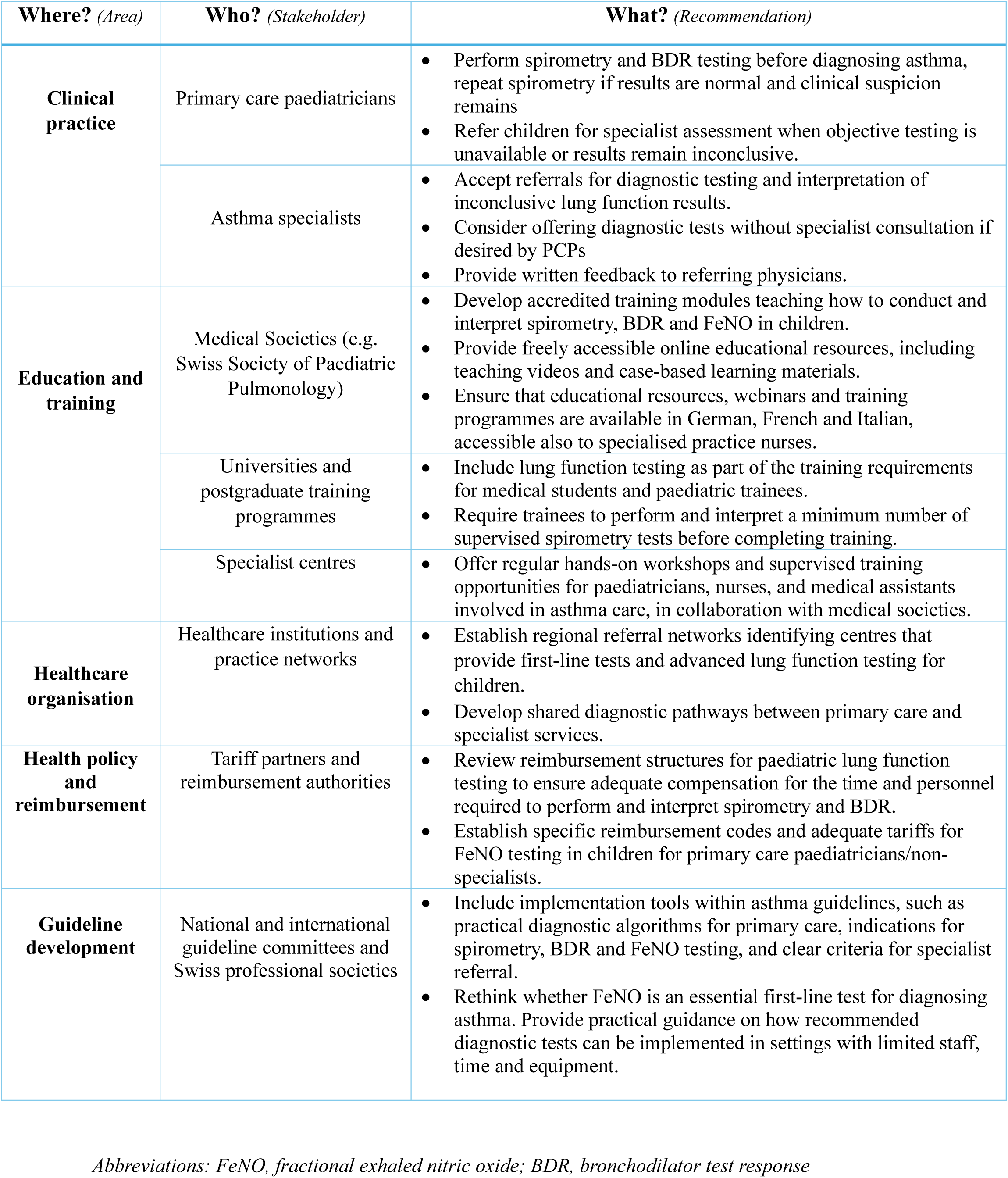
Recommendations to improve asthma diagnosis in children in Switzerland, based on results from a physician survey.

In conclusion, this study underscores the need to improve access to key diagnostic tests for the most common chronic disease in children. Implementation of evidence-based asthma diagnosis guidelines is limited not only by financial restrictions, but also by a lack of time and appropriate training in performing and interpreting these tests. Training primary care paediatricians and practice nurses in conducting, validating and interpreting lung function tests and revising reimbursement policies is essential. Without such support, guideline recommendations will remain unsuitable for application in routine clinical practice.

## DATA AVAILABILITY

Data is available upon reasonable request by contacting Claudia E Kuehni.

## AUTHORS’ ROLES

BGB, SG, MG, and CK made substantial contributions to the study design.

BGB, SG, EG, AM, NR, OS, MG, and CK made substantial contributions to the study concept and were involved in the design of the questionnaire.

BGB, MS, LL, MG, and CK made substantial contribution to the interpretation of the data. BGB analysed data and drafted the manuscript.

BGB, MS, SG, LL, EG, AM, NR, OS, MG, and CK critically revised and approved the manuscript.

## CONFLICT OF INTEREST

BGB, MS, LL, SG, OS, MG and CK have nothing to disclose. AM reports personal fees and grants from Vertex—all outside the submitted work. NR reports personal fees from AM Pharma, Schwabe Pharma, Vertex, and Sanofi—all outside the submitted work. EG reports personal fees from Sanofi and Circassia—all outside the submitted work. CK, MG, AM, NG and EG were members of the ERS guideline task force and the Swiss national guideline group.

## FUNDING

Our research was funded by the Swiss National Science Foundation, Switzerland (SNSF 320030_212519)

## ACKNOWLEDGEMENTS

We thank all physicians who participated in this survey. We also thank the Swiss Paediatric Society (*pädiatrie schweiz*), the Swiss Paediatric Pulmonology Society (*Schweizerische Gesellschaft für Pädiatrische Pneumologie*), the Swiss Pulmonology Society (*Schweizerische Gesellschaft für Pneumologie*), the Swiss Society for Allergology and Immunology (*Schweizerische Gesellschaft für Allergologie und Immunolo*gie), and the Swiss Society of Family Physicians (*Haus– und Kinderärzte Schweiz*) for their support in distributing the survey to their members.

## Notes

### Author Declarations

Under Swiss legislation, we were not required to obtain ethics committee approval or informed consent because the study was addressed to physicians and did not involve health-related personal data.

