## Supplementary tables and figures for "Asthma diagnosis in school-age children: a survey among Swiss physicians"

**Table S1.** Physician questionnaire on asthma diagnosis in school-age children.

### Section 1. Physician sociodemographic and clinical practice characteristics

- Medical specialty and level of training
- Workplace setting (private practice, hospital, both)
- Proportion of time spent in clinical work
- Practice characteristics (hospital vs practice workload)
- Number of school-age children with suspected or diagnosed asthma seen per month
- Number of new asthma diagnoses made per month
- Language region
- Gender
- Age group

### Section 2. Diagnostic tests

#### Spirometry

- Availability of spirometry in the clinic/practice
- Frequency of spirometry use for suspected asthma
- Reasons for lack of access or infrequent use
- Personnel performing spirometry
- Type of spirometer used

#### Bronchodilator reversibility testing

- Use and frequency of bronchodilator reversibility testing
- Indications for testing
- Definition of a positive test (cut-offs)
- Reasons for non-use

#### Fractional exhaled nitric oxide (FeNO)

- Availability of FeNO measurement
- Frequency of use
- Reasons for non-use or infrequent use
- FeNO cut-offs used
- Type of FeNO device

#### Other diagnostic tests

- Peak expiratory flow variability
- Free running test
- Allergy testing (type and allergens tested)
- Body plethysmography
- Bronchial provocation tests (methacholine, histamine, mannitol)
- Exercise testing
- Chest X-ray
- Use of alternative medicine
- Other tests (free text)

### Section 3. Follow-up and referral practices

- Follow-up strategy after asthma diagnosis
- Use of objective tests during follow-up
- Frequency of repeated spirometry, FeNO, BDR, body plethysmography
- Referral to specialists
- Reasons for referral

**Section 4. Trial of medication for diagnostic purposes**

- Use of trial of asthma medication to support diagnosis
- Frequency of trial of medication use
- Type of medication prescribed (SABA, ICS, ICS–LABA, LTRA, oral corticosteroids)
- Duration of treatment before reassessment
- Free-text comments on trial of medication

**Section 5. Guideline use**

- Consultation of national and international asthma guidelines
- Guidelines used (Swiss, ERS, GINA, others)
- Reasons for not using guidelines
- Agreement with Swiss and ERS guidelines
- Suggestions for guideline improvement

Note: The online questionnaire consisted of 50 core questions with conditional branching. It assessed the diagnostic practices of physicians who evaluate potential asthma in school-age children (5–16 years old), including the availability and use of objective tests, follow-up strategies, referral patterns, the use of trial medication and adherence to guidelines. Conditional branching was used to tailor questions according to specialty and access to diagnostic tests.

Above, we provide a structured overview of all the questions, grouped by theme. A full version of the questionnaire is available from the authors, upon request.

**Table S2.** Barriers to access to first-line tests in primary care. (Free text responses, translated quotes).

| Category | Reasons for not having spirometry | Reasons for not having FeNO |
|---|---|---|
| **Economic reasons** | *"Too few patients to amortize a lung function device." / "Old spirometer broken, not replaced."* | *"The purchase and maintenance costs outweigh the benefit." / "Not reimbursed for paediatricians."* |
| **Referral to specialists** | *"Refer children to paediatric pulmonologist for lung function testing." / "University hospital nearby does the tests."* | *"I refer to paediatric pulmonologist who performs FeNO." / "Nearby pneumology clinic performs the test."* |
| **No perceived need for asthma diagnosis** | *"Clinical exam sufficient for asthma diagnosis." / "Simple asthma can be diagnosed clinically, complex cases referred."* | *"Asthma diagnosis is possible with anamnesis and spirometry." / "FeNO is more useful for follow-up, not initial diagnosis."* |
| **Limited training or experience** | *"Never thought about using it." / "Not all colleagues trained in interpretation."/ "No experience with spirometry"* | *"Not trained in FeNO measurement or interpretation." / "I have never thought about this test before."* |
| **Organisational barriers** | *"I work in emergency." / "Not my practice, owner decides." / "Device expected when colleague retires."* | *"I work in emergency, no time for such tests." / "Not my decision, I'm employed." / "Small hospital, not relevant."* |
| **Positive attitude but barriers** | *"Acquisition under discussion." / "Plan to buy after training course." / "Goal for the future."* | *"I would like to have the device, but not financially viable." / "Currently under evaluation." / "Maybe if devices get cheaper."* |

**Table S3.** Determinants of access to spirometry among primary care paediatricians, based on univariable and multivariable logistic regression analyses (n = 419)

| | | | | | | |
|---|---|---|---|---|---|---|
| | **French/Italian region** | **30/155 (19%)** | **0.11 (0.06–0.18)** | **<0.001** | **0.09 (0.05–0.15)** | **<0.001** |
| **Gender** | | | | | | |
| | Female | 150/313 (48%) | 1.00 | — | — | — |
| | Male | 66/99 (37%) | 1.86 (1.17–2.99) | 0.009 | 1.51(0.81–2.83) | 0.192 |
| **Age group** | | | | | | |
| | <40 years | 47/93 (51%) | 1.00 | — | — | — |
| | 40–49 years | 78/153 (51%) | 0.92 (0.54–1.55) | 0.744 | 0.74 (0.39–1.38) | 0.341 |
| | 50–59 years | 57/108 (53%) | 0.99 (0.56–1.73) | 0.960 | 0.63 (0.31–1.26) | 0.197 |
| | ≥60 years | 34/57 (60%) | 1.75 (0.89–3.48) | 0.106 | 1.23 (0.53–2.91) | 0.636 |
| **Work percentage** | | | | | | |
| | Part-time | 155/321 (48%) | 1.00 | — | — | — |
| | Full-time | 63/98 (64%) | 2.04 (1.28–3.29) | 0.003 | 2.04 (1.11–3.82) | 0.024 |
| **Number of children with asthma seen per month** | | | | | | |
| | 1-5 children | 120/245 (49%) | 1.00 | — | — | — |
| | ≥6 children | 98/174 (56%) | 1.30 (0.87–1.93) | 0.201 | 1.17 (0.70–1.96) | 0.539 |
| **Number of new asthma diagnoses per month** | | | | | | |
| | None | 39/86 (45%) | 1.00 | — | — | — |
| | ≥ 1 child | 179/333 (54%) | 1.33 (0.82–2.19) | 0.249 | 1.30 (0.71–2.39) | 0.397 |
| **Swiss 2024 guideline** | | | | | | |
| | No use | 50/124 (40%) | 1.00 | — | — | — |
| | Use | 168/295 (60%) | 1.77(1.14–2.76) | 0.011 | 1.71 (1.01–2.92) | 0.048 |
| **ERS 2021 guideline** | | | | | | |
| | No use | 198/382 (52%) | 1.00 | — | — | — |
| | Use | 20/37 (54%) | 1.00 (0.50–1.99) | 0.992 | 0.84 (0.38–1.88) | 0.676 |
| **GINA 2023 guideline** | | | | | | |
| | No use | 122/226 (54%) | 1.00 | — | — | — |
| | Use | 106/193 (55%) | 1.29 (0.87–1.91) | 0.203 | 1.54 (0.95–2.52) | 0.080 |

*Note: OR = Odds Ratio; CI = Confidence Interval.*

*Results are from univariable and multivariable logistic regression analyses. Multivariable models were adjusted for gender, age group, language region, clinical workload (full-time vs part-time), number of children with asthma seen per month, number of new asthma diagnoses per month, and consultation of clinical guidelines.*

Reference categories: German region, female, <40 years, part-time work (<80%), 1–5 children with asthma seen per month, no new asthma diagnoses per month, and no consultation of Swiss 2024, ERS 2021, or GINA 2023 guidelines. Odds ratios >1 indicate higher odds of spirometry access compared with the reference category.

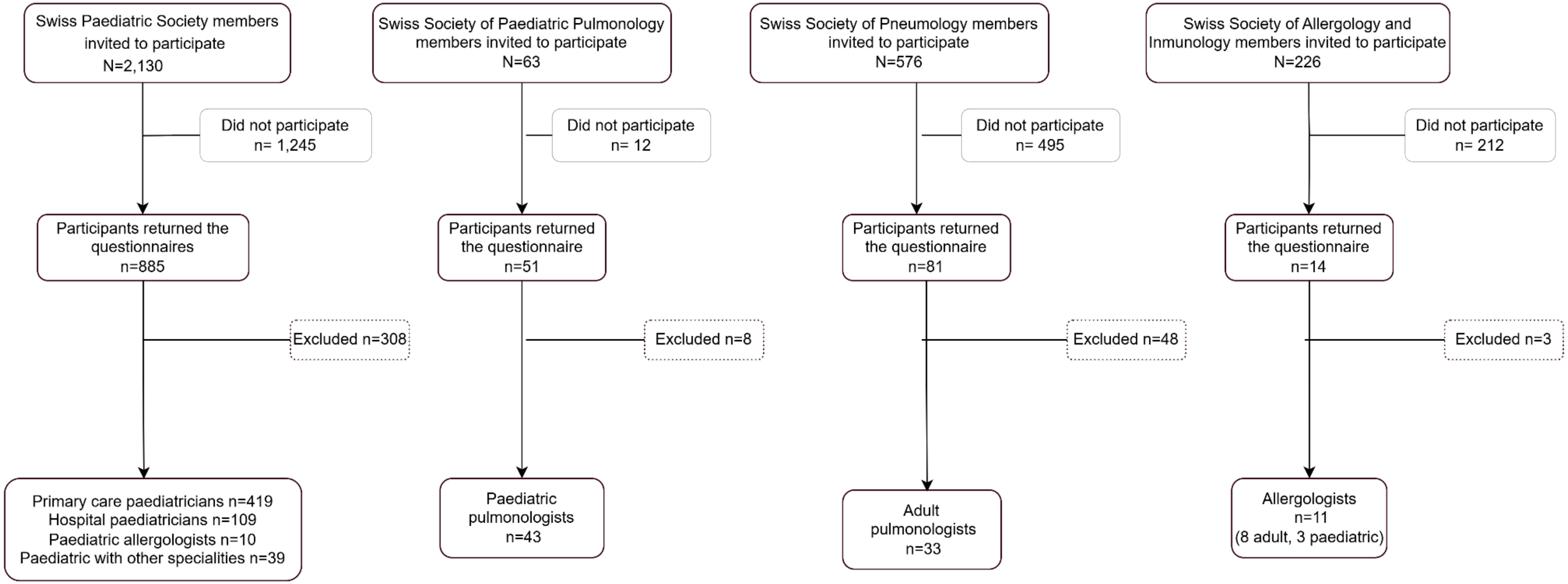


**Figure S1.** Survey invitation, response and final inclusion by Swiss medical society.

Note: Twenty-two paediatricians were members of both societies of the Swiss Paediatric Society and the Swiss Society of Paediatric Pulmonology. Each participant completed the survey only once. As they were paediatric pulmonologists, they were included in the response rate calculation for the Swiss Society of Paediatric Pulmonology.

Excluded respondents were as follows: Swiss Paediatric Society: 308 (57 did not see any children, 68 did not see children with asthma, 17 were retired, 166 did not fully complete the questionnaire). Swiss Society of Paediatric Pulmonology: 8 (2 retired, 6 did not fully complete the questionnaire). Swiss Society of Pulmonology: 48 (1 did not see any children, 26 did not see children with asthma, 4 retired, 17 did not fully complete the questionnaire). Swiss Society of Allergology and Immunology: 3 (all did not see children with asthma). General practitioners were excluded from the analysis due to the very low response rate: 24 (8 did not see children with asthma, 2 retired, 10 did not fully complete the questionnaire, 4 complete the questionnaire)

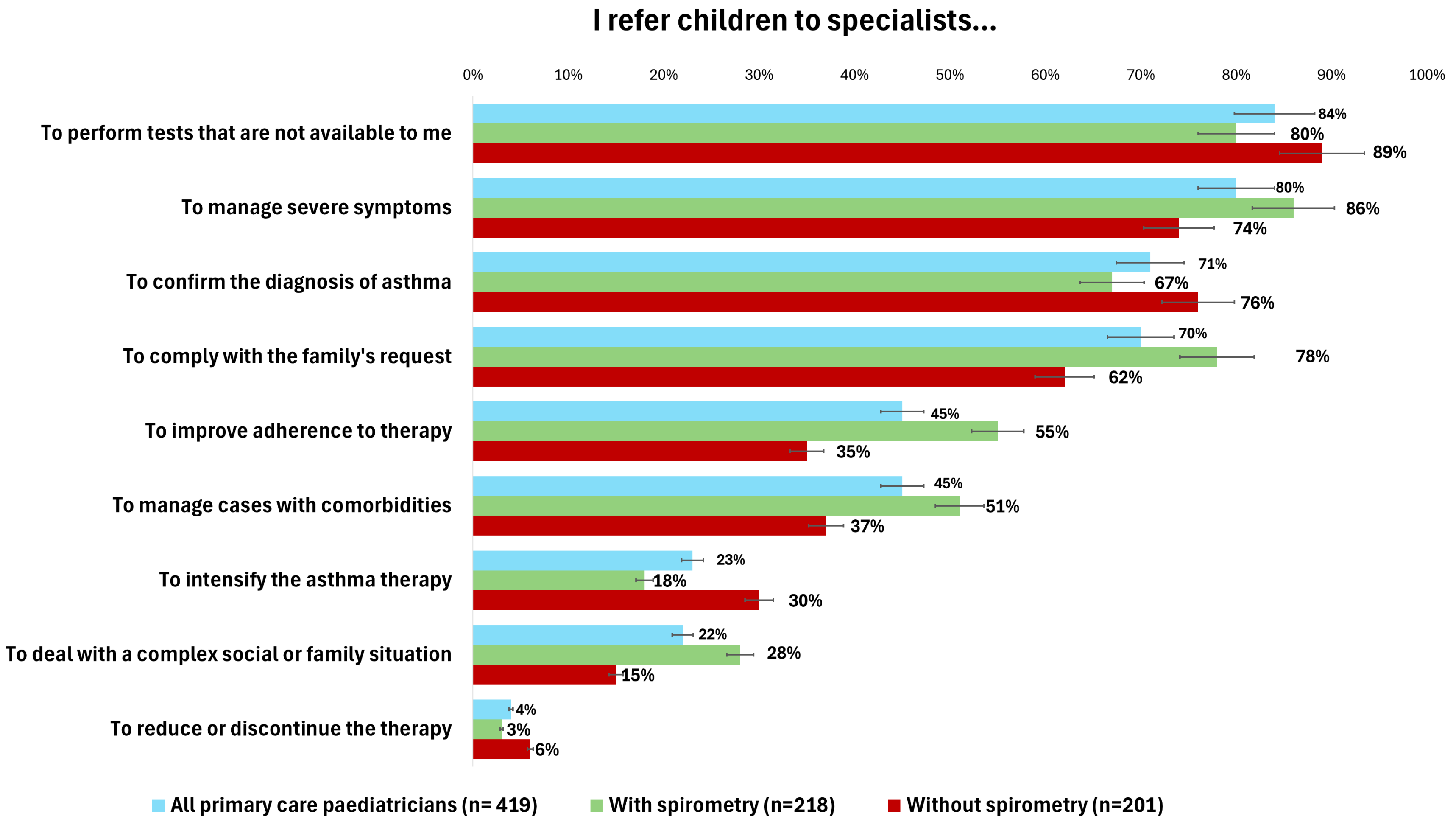


**Figure S2.** Referral reasons of children with suspected asthma to a specialist reported by PCPs (n = 419), overall and by access to spirometry.

Note: Bars show percentages with 95% CIs; numbers indicate %. Multiple choice question.
